# GIS-based spatial modeling to identify factors affecting COVID-19 incidence rates in Bangladesh

**DOI:** 10.1101/2020.08.16.20175976

**Authors:** Md. Hamidur Rahman, Niaz Mahmud Zafri, Fajle Rabbi Ashik, Md Waliullah

## Abstract

The outbreak of the COVID-19 pandemic is an unprecedented shock throughout the world which leads to generate a massive social, human, and economic crisis. However, there is a lack of research on geographic modeling of COVID-19 as well as identification of contributory factors affecting the COVID-19 in the context of developing countries. To fulfill the gap, this study aimed to identify the potential factors affecting the COVID-19 incidence rates at the district-level in Bangladesh using spatial regression model (SRM). Therefore, data related to 32 demographic, economic, weather, built environment, health, and facilities related factors were collected and analyzed to explain the spatial variability of this disease incidence. Three global (Ordinary least squares (OLS), spatial lag model (SLM) and spatial error model (SEM)) and one local (geographically weighted regression (GWR)) SRMs were developed in this study. The results of the models showed that four factors significantly affected the COVID-19 incidence rates in Bangladesh. Those four factors are urban population percentage, monthly consumption, number of health workers, and distance from the capital. Among the four developed models, the GWR model performed the best in explaining the variation of COVID-19 incidence rates across Bangladesh with a *R^2^* value of 78.6%. Findings from this research offer a better insight into the COVID-19 situation and would help to develop policies aimed to prevent the future epidemic crisis.

## Introduction

Coronavirus disease (COVID-19) is a very infectious disease caused by the SARS-CoV-2 virus. The first human cases of COVID-19 were reported in Wuhan City, China, in December 2019 (WHO, 2020a). Within some weeks, the outbreak of COVID-19 spread globally; as a result, the World Health Organization (WHO) declared this as a public health emergency of international concern (PHEIC) on 30 January 2020 (WHO, 2020b) and a pandemic on 11 March 2020 (WHO, 2020c). As of 2:46 pm CEST, 9 August 2020, there have been 19,462,112 reported cases resulting 722,285 deaths (WHO, 2020d). According to the World Bank, COVID-19 has triggered a global crisis like no other, as well as this is leading to the deepest global recession since the Second World War. The baseline forecast envisions a 5.2% contraction in the global GDP in 2020— the deepest global recession in the previous eight decades, despite unprecedented policy support (WorldBank, 2020). About 1.6 billion informal workers having little to no savings and no access to social protection, lost 60% of their income. This pandemic will push 40–60 million people into extreme poverty (UN, 2020).

In Bangladesh, on 8 March 2020, the first three coronavirus cases were confirmed by the Institute of Epidemiology Disease Control and Research (IECDR, 2020). Affected persons were returned from Italy for joining their family in their native place (Daily Sun, 2020). On 18 March, Bangladesh reported its first coronavirus death (NewAge, 2020). Thereafter, the virus transmitted from capital city, Dhaka, to other major administrative areas of Bangladesh rapidly. To hold the spread of the disease, the government declared nationwide ‘lockdown’ form 26 March to 30 May (IMF, 2020). As of 9 August 2020, 257,600 cases have been reported resulting in 3,399 death (**Figure 1**) (IECDR, 2020). Like most of the other developing nations, the outbreak of the COVID-19 pandemic is an unprecedented shock to Bangladesh in terms of social, human, and economic crisis.

**Figure 1:**
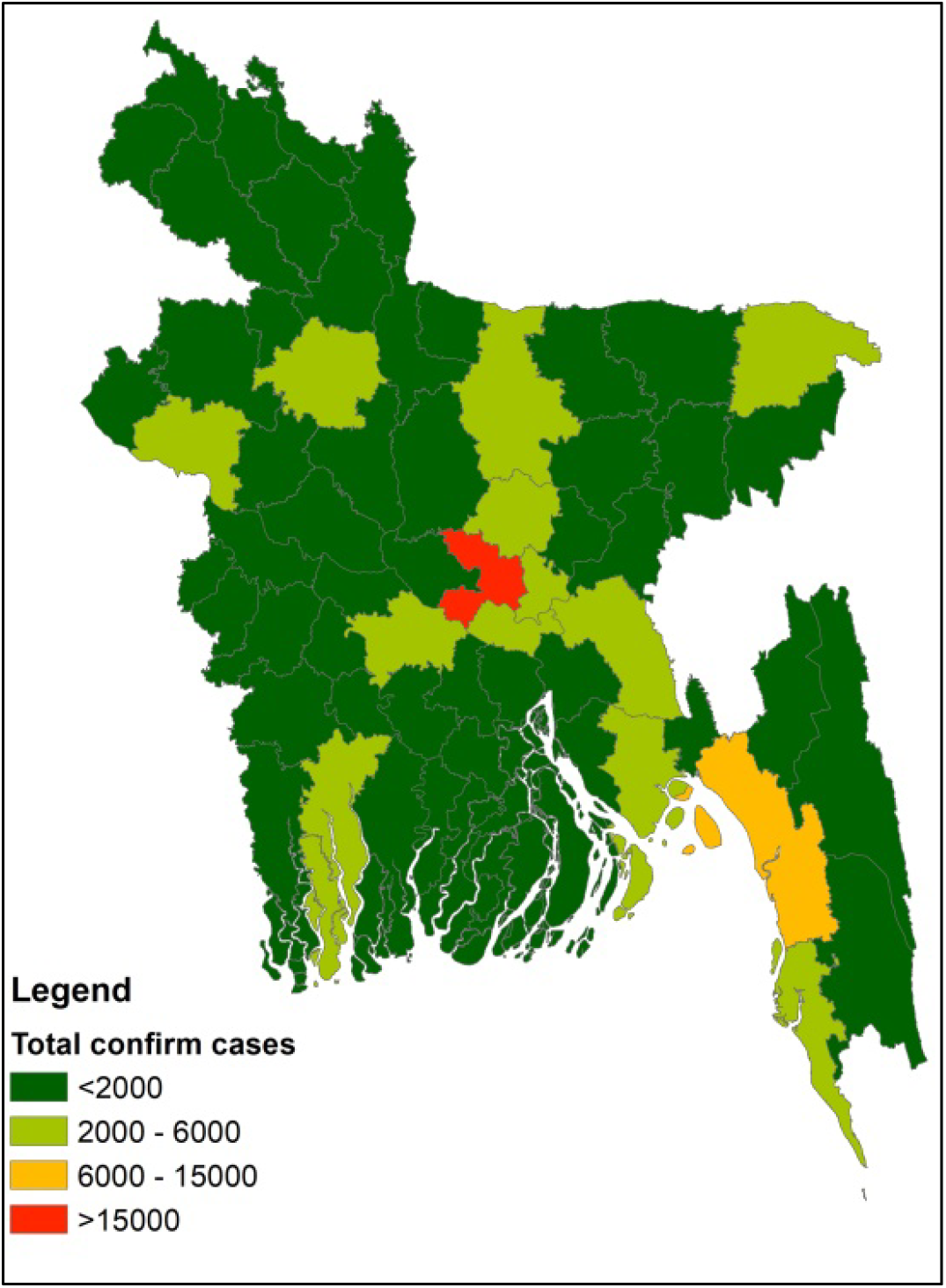
Number of confirmed COVID-19 cases by district

Disadvantaged demographic pattern, economic, environmental, and health condition have been established as potential determinants of infectious diseases in general (Khalatbari-Soltani, Cumming, Delpierre, & Kelly-Irving, 2020). A brief overview of the literature suggested that socio-economic and demographic factors had a significant association with the efficacy of vaccination for Tuberculosis in Germany (Bluhm & Pinkovskiy, 2020), for Salmonella infections in USA (Varga et al., 2013), for HIV (Bärnighausen, Hosegood, Timaeus, & Newell, 2007), for Pertussis in Australia (Huang et al., 2017), for Pneumonia and Influenza (Farr, Bartlett, Wadsworth, & Miller, 2000). To be specific, areas with low population density and population with older age group (> = 65+ years) had a high percentage of H1N1 deaths (Ponnambalam, Samavedham, Lee, & Ho, 2012). In addition, population density and medical staff density had a positive influence on SARS (Fang et al., 2009), educational status and living condition had a positive impact on H1N1 influenza rate (Lowcock, Rosella, Foisy, McGeer, & Crowcroft, 2012), economic activity type and educational institutions location had an impact on the incidence of Hand-foot-mouth disease in China (Huang et al., 2017), household-level sanitation infrastructure had impact on avian influenza in Vietnam (Spencer et al.,2020), and the urban land area had direct impact on influenza mortality rate in 1918–20 (Mamelund, 2011). These findings suggest that similar results can be obtainable for the newly emerged coronavirus disease. Identification of possible demographic, economic, weather, built environment, health, and service facilities related factors are crucial at each phase of the epidemic to effectively interrupt human to human transmission chains as well as preventing further spread through appropriate interventions (Khalatbari-Soltani et al., 2020).

However, the impact of these indicators on the COVID-19 transmission and their magnitude is yet under scrutiny and investigation. So far, there has been attempt to identify contributing factors, i.e., social (Corburn et al., 2020; Khalatbari-Soltani et al., 2020), economic (Atkeson, 2020), health (Zhao et al., 2020), demographic (Hamidi, Sabouri, & Ewing, 2020; Sannigrahi, Pilla, Basu, & Basu, 2020), environmental (Liu et al., 2020; Mollalo, Vahedi, & Rivera, 2020; Qi et al., 2020) which might affect the spread, morbidity rate, and mortality rate of the COVID-19 disease. Though several studies conducted for identifying factors influencing the COVID-19 pandemic, more studies need to be conducted in different contexts as well as considering a wide variety of factors to get a complete scenario. Country contexts are very important as developing countries have major issues like high population, lack of local infrastructure, social equality, variety of employment, and inefficacy of public health measures, which might bring disparate outcome in terms of virus incidence rates compared to developed countries (Khalatbari-Soltani et al., 2020). Bangladesh, a highly dense developing country, is a perfect study unit for understanding the impact of contributing determinants on the COVID-19 outbreak. In addition, a large number of factors (i.e., built environment, service facilities related factors and others) were not considered in the previous studies, which need to be explored to get a comprehensive knowledge about the pandemic.

Geographic information system (GIS) is an essential tool which can assist in the process of combating a pandemic as well as improve the quality of care through examining the spatial distribution of infectious diseases (Lovett et al., 2014; Mollalo, Mao, Rashidi, & Glass, 2019; Mollalo et al., 2018). A limited number of literatures have been published since the outbreak of COVID-19, where GIS has been used for identification of early spread of COVID-19 (Guan et al., 2020), distribution of active cases (Chen et al., 2020), mapping (Dong, Du, & Gardner, 2020), comparing spatio-temporal patterns (Zhang, Rao, Wu, Huang, & Dai, 2020), the effectiveness of containment measure (Orea & Álvarez, 2020), and identification of virus risk factors(Mollalo et al., 2020). The use of geospatial and statistical tools is critical to explore the association between COVID-19 incidence and its contributing factors as it is a process which occurs in geographical space (Gross et al., 2020; Mollalo et al., 2019). Traditional statistical approaches used in epidemiological studies, i.e., factor analysis (Meigs, 2000), principal component analysis (Varraso et al., 2012), cluster analysis (Merlo et al., 2006), regression analysis (Blyth, Kincaid, Craigen, & Bennet, 2001) fail to take account the spatial dependency and autocorrelation in parameters estimation. To address this functional lag, spatial regression model (SRM), i.e., spatial lag model (SLM), spatial error model (SEM), and geographically weighted regression (GWR) has been widely used in epidemiological studies (Sannigrahi et al., 2020). For the COVID-19 pandemic, until now, there are few studies available which used SRM to find out the determinants of incidence rates of this virus. Therefore, this study aimed to identify potential demographic, social, economic, weather, built environment, health, and facilities related determinants of the COVID-19 incidence rates at district-level across Bangladesh using SRMs. The objective of the study was: (1) to identify the key explanatory factors which have significant impact on the COVID-19 incidence rates and (2) to quantify the spatial relationships between contributing factors and the COVID-19 incidence rates. Findings from this study would help the developing countries to prepare equitable public health prevention measures and guidelines for any future pandemic situation.

## 2. Data and methods

### 2.1 Data collection and preparation

Institute of Epidemiology, Disease Control and Research (IEDCR) has been monitoring the spread of the pandemic and updating the database of COVID-19 daily basis at district and city level across Bangladesh. For this study, number of positive COVID-19 cases data at district-level across Bangladesh were considered from March 8, 2020 (first known cases in Bangladesh) to July 28, 2020, retrieved from the IECDR data portal (https://iedcr.gov.bd/). COVID-19 cases per 10,000 population per district was considered as the dependent variable for modeling and interpretation purpose and termed as COVID-19 incidence rates in this study.

A total of 32 demographic, economic, weather, built environment, health facilities, and community facilities related factors were considered as explanatory variables for the model development. Most of the data of relevant factors were compiled from different database of the government of Bangladesh, i.e., Bangladesh Bureau of Statistics (BBS) (http://www.bbs.gov.bd/) which is the centralized official bureau in Bangladesh for collecting statistics on demography, economy, and other facts of the country; Bangladesh Metrological Department (BMD) (http://www.bmd.gov.bd/) which is the national meteorological organization of Bangladesh. Few factors were derived through spatial interpolation and analysis using ArcGIS Pro 2.4 software. Detail description of the variables and their sources are showed in **Table 1**. After data collection and preparation, district-level information was joined with the corresponding district in ArcGIS.

**Table 1:**
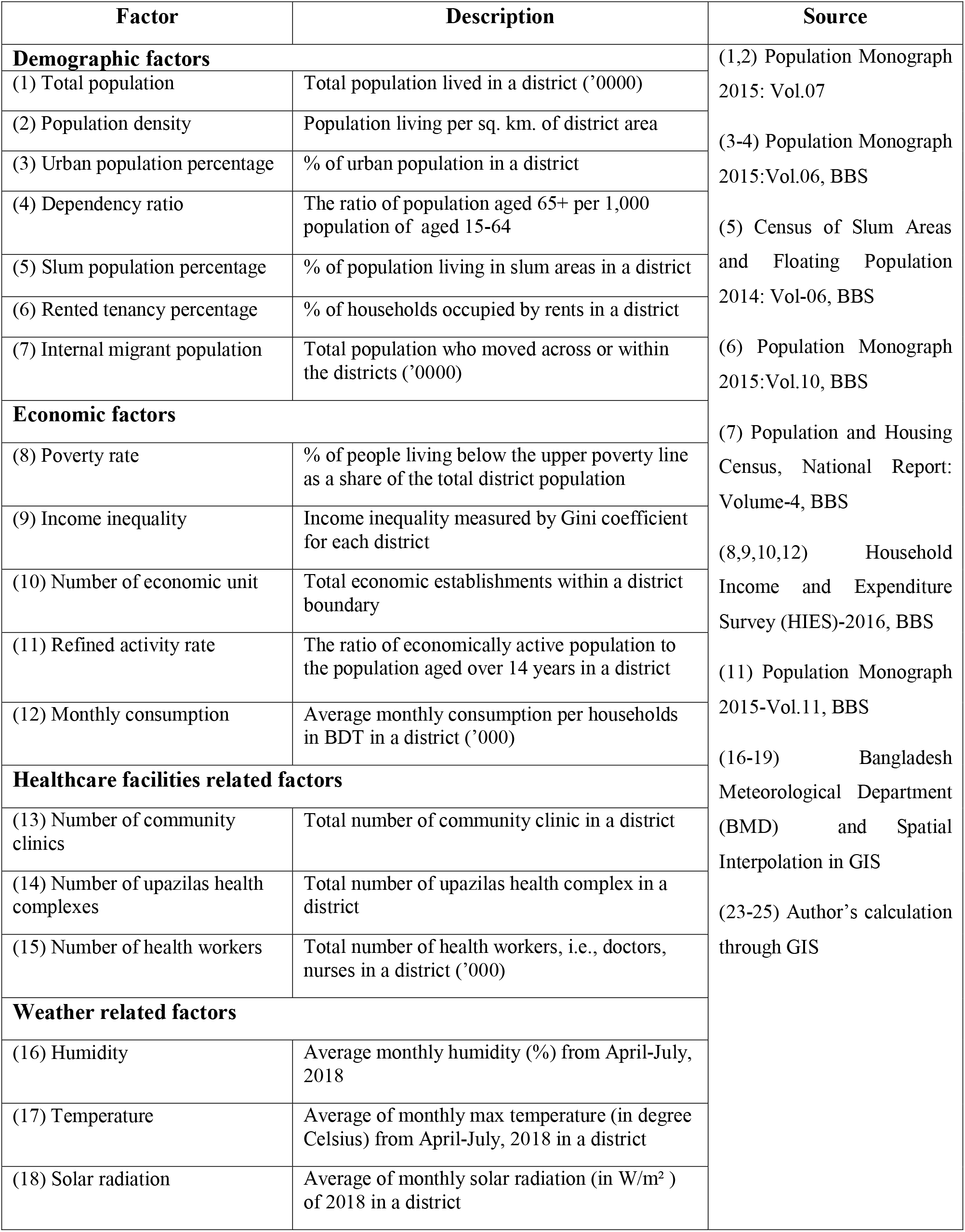
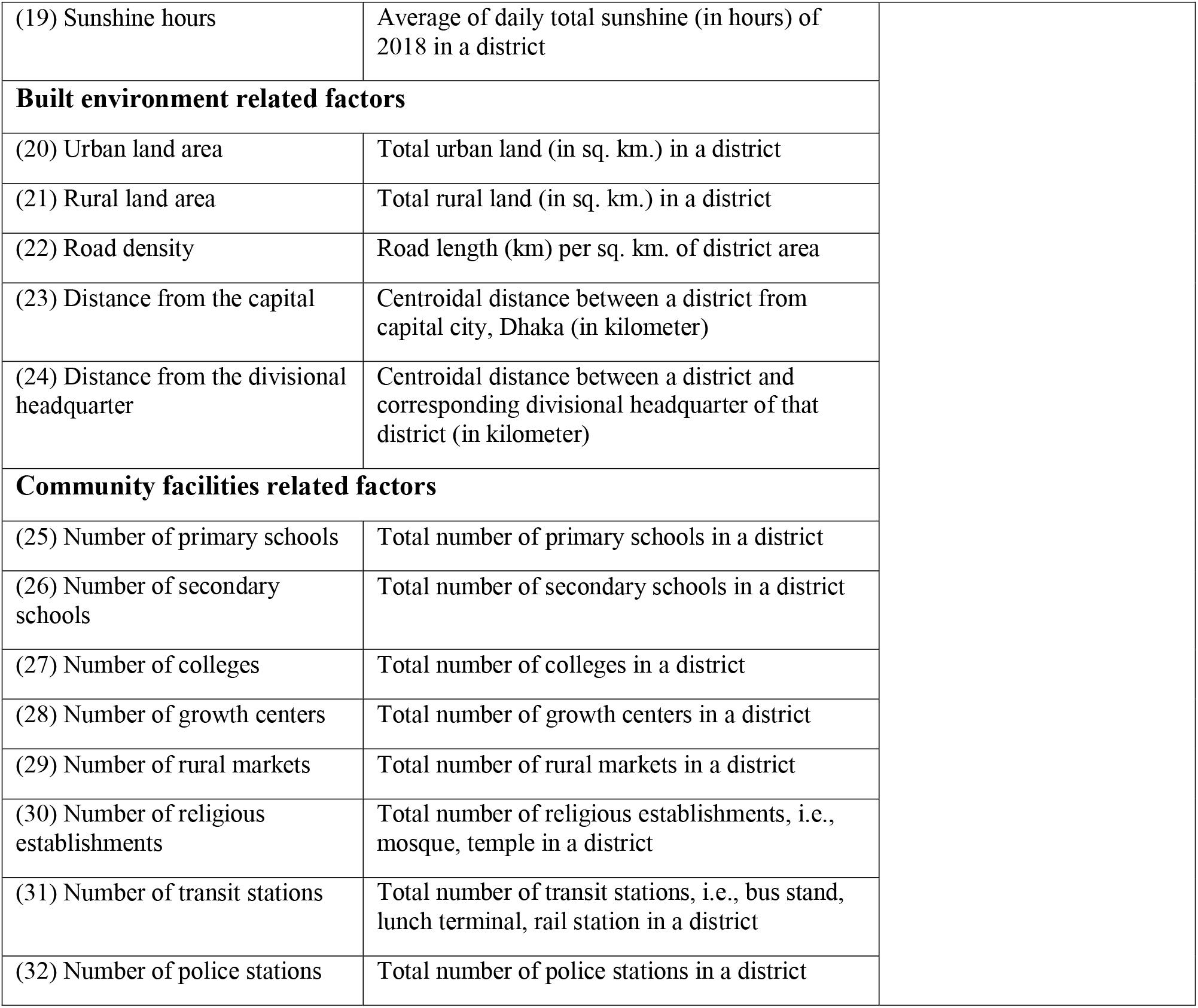
Collected factors used in this study together with definitions and sources

Data of considering explanatory variables of interest in this study are inherently spatial in nature. Hence, it is plausible that data would be positively spatially dependent which means areas located nearby tend to be more similar than those separated by great distances (Wheeler & Páez, 2010). Therefore, both local and global regression models were used to determine how well they could explain the variation of COVID-19 incidence rates at district-level of Bangladesh. The global models included OLS, SLM, SEM; whereas, local model included GWR.

### 2.2 Global models

#### 2.2.1 Ordinary least squares (OLS)

Ordinary least squares regression is a statistical method that estimates the relationship between a set of explanatory or independent variables and a dependent variable with the fundamental assumption of homogeneity and spatial non-variability (Oshan, Li, Kang, Wolf, & Fotheringham, 2019; Ward & Gleditsch, 2018). The form of the OLS model used in this research:

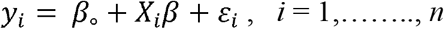

Here,*y*_i_ is the COVID-19 incidence rates in district i,*β_o_* is the intercept, *X_i_* is the vector of selected explanatory variables, *β* is the vector of regression coefficients, and *ε_i_* is a random error term.

The fundamental function of OLS is to optimize the regression coefficients (β) by minimizing the sum of the squares in the difference between the observed and predicted values of the dependent variable configured as a straight line (Anselin & Arribas-Bel, 2013; Mollalo et al., 2020; Oshan et al., 2019). Two implicit assumptions of OLS method: (1) the observations are independent and homogenous across the study area; (2) error terms are not correlated (Anselin & Arribas-Bel, 2013; Oshan et al., 2019), lead to bias in regression coefficient estimation when the errors are heterogeneous and spatially correlated (Goodchild, Parks, & Steyaert, 1993; Yang & Jin, 2010). In the case of COVID-19 incidence, possible explanatory variables used in this research are spatially correlated, which denote that an explanatory variable could be influenced by another explanatory variable in the neighboring district (as observed from the SEM, SLM results). As OLS failed to capture these interactions, we used SEM and SLM, which take spatial dependence into account, but model it differently (Anselin, 2003; Ward & Gleditsch, 2018).

#### 2.2.2 Spatial lag model (SLM)

The SLM can accommodate the spatial dependency between the dependent variable and explanatory variables by incorporating a “spatially-lagged dependent variable” in the regression model (Anselin, 2003; Mollalo et al., 2019; Mollalo et al., 2020; Ward & Gleditsch, 2018). SLM is denoted as-

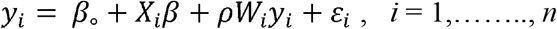

Where: at district *i, y_i_* = COVID-19 incidence rates in district *i; β_o_* = intercept; *X_i_* = matrix of observations of selected explanatory variables; *ρ* = spatial autoregressive parameter which measures the intensity of spatial interdependency (Rho), and *W_i_* = vector of spatial weights which specifies how features are related to each other/ The weight matrix (W) specifies how the neighbors at district *i* and connects one independent variable to the explanatory variables at that location (Anselin & Arribas-Bel, 2013). A spatial lag is a variable that averages the neighboring values of a location (Sannigrahi et al., 2020). The SLM accounts for autocorrelation in the model with the weight matrix.

#### 2.2.3 Spatial error model (SEM)

The SEM model assumes that OLS residuals or error terms have spatial dependence or spatially correlated (Anselin & Arribas-Bel, 2013). Hence, residuals are decomposed into two terms-error terms and a random error term (Mollalo et al., 2020; Sannigrahi et al., 2020), where the general form of the model is:

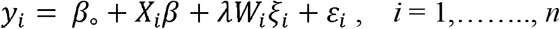

Where: at district *i, ξ_i_* = indicates the spatial component of the error; *λ* = indicates the λ level of correlation between these components (Lamda); *ε_i_* = spatially uncorrelated error term/ independent identically distributed errors; *W_i_* = spatial weights matrix; *W_i_ξ_i_* = the extent to which the spatial component of the errors is correlated with one another for nearby observations. The SEM accounts autocorrelation in the error with the weights matrix.

### 2.3 Local model

#### 2.3.1 Geographically weighted regression (GWR)

The GWR is a local form of linear regression as it is used to model the spatially varying association between a dependent and independent variable (Brunsdon, Fotheringham, & Charlton, 1996). The global regression model, i.e., OLS, SEM, and SLM method have one prominent limitation while it is applied to a spatial dataset (Deilami, Kamruzzaman, & Hayes, 2016; Mollalo et al., 2020; Sannigrahi et al., 2020). This model cannot account for a spatial nonstationarity issue which explains the relationship between the dependent and independent variables might vary over space (Brunsdon et al., 1996; Sannigrahi et al., 2020). Thus, global regression estimates parameters that are average of the entire area of interest rather than specific locations within an area (Deilami & Kamruzzaman, 2017; Deilami et al., 2016). The GWR model overcomes this limitation by increasing the local effectiveness of the model by incorporating geographic context where parameters are derived for each location separately (Oshan et al., 2019). Mathematically, the general form of the GWR model is (Deilami et al., 2016; Mollalo et al., 2020):

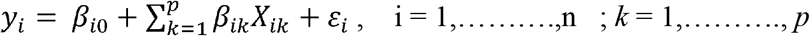

Where: at district *i, K* = independent variable within each district which varies from variable 1 to variable *p; X_ik_* = value of *K^th^* independent variable; *λ_ik_* = local regression coefficient for *K^th^* independent variable; *λ_i0_* = intercept parameter, and *ε_i_* =random disturbance.

The GWR model is sensitive to kernel type and kernel bandwidth which together define a moving window to determine the parameters, i.e., *R^2^*, coefficients in the mode (Deilami et al., 2016; McMillen, 2004). Among two types of the kernel, a fixed kernel uses a constant distance while the adaptive kernel uses the varying distance from place to place. In terms of bandwidth, Akaike Information Criterion (AICc), Cross-Validation (CV), and bandwidth-parameter are mostly used (Fotheringham, Brunsdon, & Charlton, 2003; Páez, Farber, & Wheeler, 2011). The rationale behind using each of these options is discussed in (Fotheringham et al. (2003)) and not presented here.

### 2.4 Models development

To identify the potential explanatory variables for developing multivariate models from the collected data of a large number of factors, univariate analysis was conducted through OLS model to identify the impact of each variable on the COVID-19 incidence rates individually. OLS models were developed in ArcGIS software. Factors that were found to be insignificant in this analysis were not considered further as explanatory variables. Then, Pearson’s correlation analysis was conducted to examine the correlations between the factors which were significant in univariate analysis. Subsequently, several highly related factors were eliminated to avoid multicollinearity in the models. The rest of the factors were considered as explanatory variables and inputted for developing an overall global multivariate OLS model. The final OLS model was developed through trial and error procedure. For detecting multicollinearity in the model, Variance Inflation Factor (VIF) was used. After that, SLM, SEM, and GWR were developed using the explanatory variables of the final OLS model. Two global models (SLM and SEM) were developed in GeoDa 1.14 software (geodacenter.github.io). Based on first-order Queen’s contiguity, the weight matrix was generated for the SLM and SEM. On the other hand, local model (GWR) was run in ArcGIS software. *Adaptive* kernel type and *AICc* bandwidth were selected to run the GWR model. Finally, The *R^2^* and *AICc* values of the four developed models were used to compare the performances of the models in explaining COVID-19 incidence rates across Bangladesh.

## 3. Results

Results of the univariate analysis are presented in **Table 2**. Among the 32 considering factors, 17 factors were found statistically significant and considered as explanatory variables for developing a multivariate model later. All the demographic factors were found significant; whereas, not a single variable was found significant from weather related factors. From **Table 2**, it is also clear that demographic and health related most of the factors have comparatively higher *R^2^* values which mean these factors could explain a good portion of the variation in the COVID-19 incidence rates across Bangladesh. On the other hand, relatively lower *R^2^* was found for other factors. Therefore, variation in COVID-19 incidence rates could mostly describe by demographic and health related factors.

**Table 2:** Results of univariate analysis

| **Factor** | **Intercept** | **Coefficient** | **Std. Error** | **p-value** | **R<sup>2</sup>** |
| --- | --- | --- | --- | --- | --- |
| **Demographic factors** |  |  |  |  |  |
| Total population | 2.492 | 0.023 | 0.004 | 0.000* | 0.366 |
| Population density | 2.658 | 0.004 | 0.001 | 0.000* | 0.507 |
| Urban population percentage | -0.973 | 0.476 | 0.054 | 0.000* | 0.561 |
| Dependency ratio | 18.684 | -0.163 | 0.074 | 0.032** | 0.072 |
| Slum population percentage | 5.035 | 2.791 | 0.473 | 0.000* | 0.359 |
| Rented tenancy percentage | 4.348 | 0.462 | 0.046 | 0.000* | 0.519 |
| Internal migrant population | 6.166 | 0.069 | 0.008 | 0.000* | 0.529 |
| **Economic factors** |  |  |  |  |  |
| Poverty rate | 12.294 | -0.168 | 0.050 | 0.001* | 0.153 |
| Income inequality | 11.316 | -13.007 | 21.749 | 0.552 | 0.006 |
| Number of economic unit | 2.675 | 0.040 | 0.007 | 0.000* | 0.344 |
| Refined activity rate | -15.887 | 0.803 | 0.191 | 0.000* | 0.221 |
| Monthly consumption | -5.766 | 0.928 | 0.189 | 0.000* | 0.279 |
| **Healthcare facilities related factors** |  |  |  |  |  |
| Number of community clinics | 9.453 | -0.013 | 0.015 | 0.389 | 0.012 |
| Number of upazilas health complexes | 8.295 | -0.029 | 0.038 | 0.454 | 0.009 |
| Number of health workers | 5.068 | 1.253 | 0.149 | 0.000* | 0.533 |
| **Weather related factors** |  |  |  |  |  |
| Humidity | -31.714 | 0.497 | 0.379 | 0.195 | 0.027 |
| Temperature | -0.740 | 0.254 | 1.151 | 0.826 | 0.001 |
| Solar radiation | -34.985 | 0.258 | 0.160 | 0.112 | 0.040 |
| Sunshine hours | -25.309 | 5.658 | 7.986 | 0.481 | 0.008 |
| **Built environment related factors** |  |  |  |  |  |
| Urban land area | 4.928 | 0.020 | 0.010 | 0.049** | 0.060 |
| Rural land area | 7.674 | -0.001 | 0.001 | 0.986 | 0.000 |
| Road density | 6.464 | 0.482 | 0.615 | 0.435 | 0.010 |
| Distance from the capital | 12.315 | -0.031 | 0.010 | 0.004* | 0.127 |
| Distance from the divisional headquarter | 11.225 | -0.049 | 0.017 | 0.006* | 0.114 |
| **Community facilities related factors** |  |  |  |  |  |
| Number of primary schools | 5.825 | 0.002 | 0.002 | 0.356 | 0.014 |
| Number of secondary schools | 5.929 | 0.006 | 0.006 | 0.313 | 0.016 |
| Number of colleges | 6.030 | 0.036 | 0.034 | 0.284 | 0.018 |
| Number of growth centers | 5.411 | 0.070 | 0.064 | 0.274 | 0.019 |
| Number of rural markets | 8.450 | -0.003 | 0.007 | 0.642 | 0.004 |
| Number of religious establishments | 5.189 | 0.001 | 0.000 | 0.134 | 0.036 |
| Number of transit stations | 4.879 | 0.053 | 0.018 | 0.005* | 0.123 |
| Number of police stations | 4.122 | 0.162 | 0.044 | 0.001* | 0.179 |

\* significant at 99% confidence level, \*\* significant at 95% confidence level

For developing an overall global multivariate OLS model, correlation analysis was conducted to address multicollinearity issue among the explanatory variables (**Figure 2**). Several factors, had high *R^2^* and was found strongly significant in univariate analysis, were not considered for the model development to diminish multicollinearity in the model. Those factors were: total population, population density, rented tenancy percentage, internal migrant population, and number of economic unit. Several OLS models were developed through trial and error procedure using rest of the explanatory variables. Among them, a final model was selected based on having the highest *R^2^* value. This final global OLS model included only four factors. These factors are urban population percentage, monthly consumption, number of health workers, and distance from the capital (**Table 3)**. The model has relatively low multicollinearity since the highest VIF value among the four factors (VIF = 2.9) is far less than the threshold of 7.5. In addition, only distance from the capital factor was negatively associated with the COVID-19 incidence rates. This association was found positive for the rest of the three factors. This model was found to be statistically significant and have a *R^2^* value of 0.673. This value means that about 67.3% of the COVID-19 incidence rates across Bangladesh are caused by these four factors of the model. Rest of the 32.7% incidence rates are caused by unknown factors to the model and probably for the local variations which could not captured by the global OLS model.

**Figure 2:**
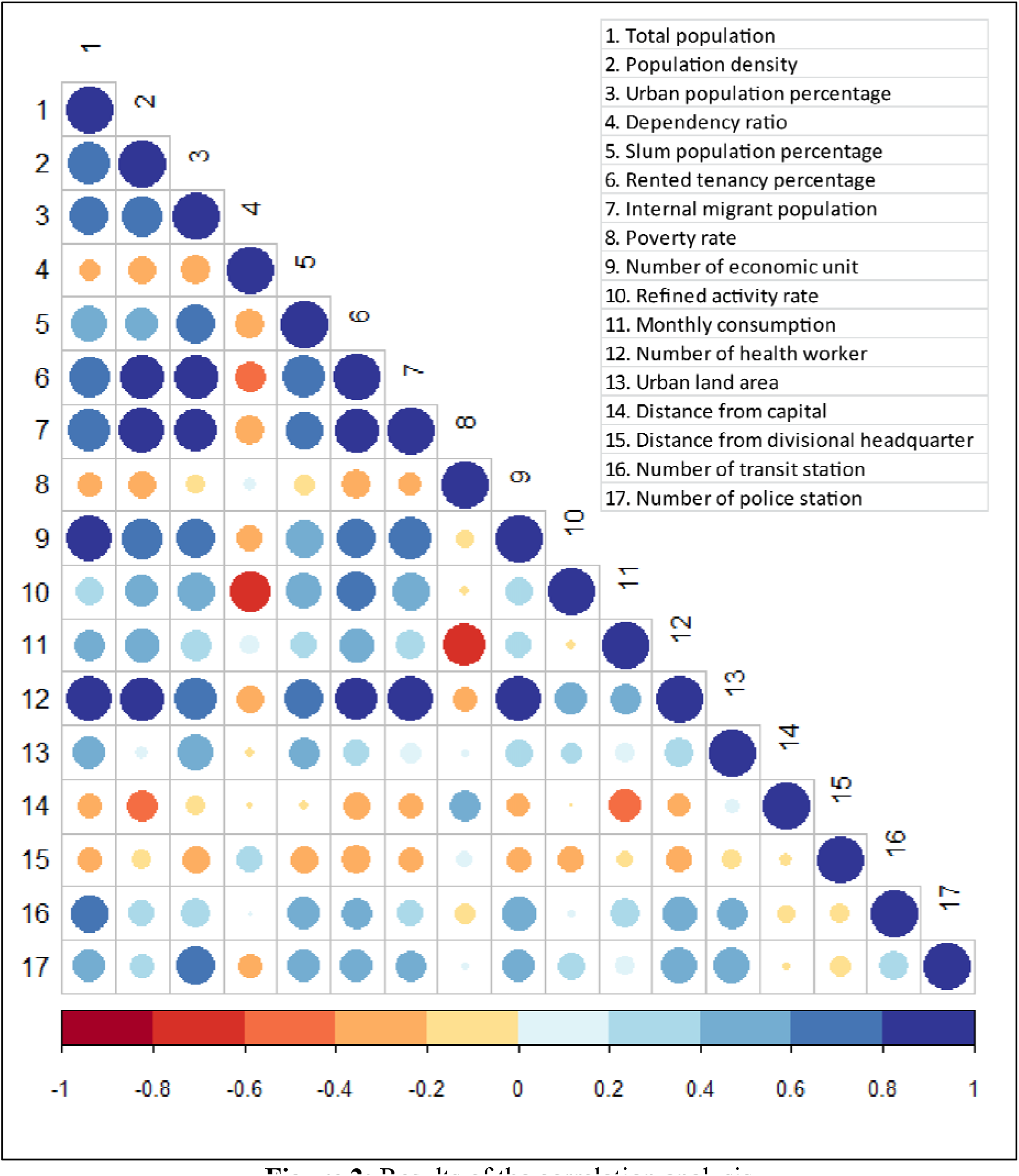
Results of the correlation analysis

**Table 3:** Results of the final multivariate OLS model

| Factors | Coefficient | Std. error | p-value | VIF |
| --- | --- | --- | --- | --- |
| Intercept | -0.966 | 2.890 | 0.738 |  |
| Urban population percentage | 0.278 | 0.080 | 0.000* | 2.857 |
| Monthly consumption | 0.314 | 0.157 | 0.046** | 1.450 |
| Number of health workers | 0.468 | 0.217 | 0.035** | 2.903 |
| Distance from the capital | -0.013 | 0.007 | 0.079*** | 1.263 |
| Model statistics: $F(4, 59) = 30.401$, $p = 0.000$, $R^2 = 0.673$ | | | | |

\* significant at 99% confidence level, \*\* significant at 95% confidence level, \*\*\* significant at 90% confidence level

To improve the performance of the overall OLS model by incorporating spatial dependence, SLM and SEM were developed (**Table 4**). Both autoregressive lag coefficients (Rho and Lamda) of the models were found to be strongly statistically significant at 99% confidence level. Both SLM and SEM have higher *R^2^* values and lower *AICc* values than the OLS model (**Table 5**). Therefore, it can be said that the SLM and SEM performed better than the OLS model. However, the performance of modeling the COVID-19 incidence rates in Bangladesh might be improved more if the model developed in local scale instead of global scale.

**Table 4:** Results of the SLM and SEM model

| Variable | Coefficient |  | Std. error |  | p-value |  |
| --- | --- | --- | --- | --- | --- | --- |
|  | SLM | SEM | SLM | SEM | SLM | SEM |
| Intercept | -2.309 | -1.558 | 2.583 | 3.251 | 0.371 | 0.631 |
| Urban population percentage | 0.237 | 0.299 | 0.074 | 0.072 | 0.000* | 0.000* |
| Monthly consumption | 0.181 | 0.318 | 0.149 | 0.164 | 0.033** | 0.037** |
| Number of health workers | 0.565 | 0.346 | 0.202 | 0.182 | 0.005* | 0.057*** |
| Distance from the capital | -0.006 | -0.011 | 0.006 | 0.009 | 0.025** | 0.015** |
| Rho | 0.356 |  | 0.113 |  | 0.001* |  |
| Lamda |  | 0.466 |  | 0.135 |  | 0.000* |

\* significant at 99% confidence level, \*\* significant at 95% confidence level, \*\*\* significant at 90% confidence level

**Table 5:** Measures of goodness-of-fit for OLS, SEM, SLM, and GWR models

| Criterion | OLS | SLM | SEM | WGR |
| --- | --- | --- | --- | --- |
| $R^2$ | 0.673 | 0.717 | 0.722 | 0.786 |
| $AICc$ | 363.94 | 355.18 | 353.63 | 340.49 |

GWR was used to model the COVID-19 incidence rates in local scale. From **Table 5**, *R^2^* value was found the highest for SEM model among the global models. This value increased from 0.722 in the SEM to 0.786 in the GWR model. Therefore, it is clear that GWR model could explain 78.6% of the variations of COVID-19 incidence rates across Bangladesh. In addition, *AICc* value was also found the lowest in the GWR model (*AICc* = 340.49) compared to the others global models, indicating GWR model is the most parsimonious model.

The spatial distributions of the coefficient values of GWR model are presented in **Figure 3**. As seen in **Figure 3**, monthly consumption and number of health workers factors demonstrated nearly similar patterns; opposite pattern was found for urban population percentage. For eastern and southern parts of Bangladesh, monthly consumption and number of health workers were found as influential factors and urban population percentage was found as weak factor in explaining the COVID-19 incidence rates. On the other hand, for the western and northern parts of the country, opposite findings were observed. Furthermore, distance from the capital was found as an influential factor in the central parts of the country. It was less influential in the south-eastern parts of the country.

**Figure 3:**
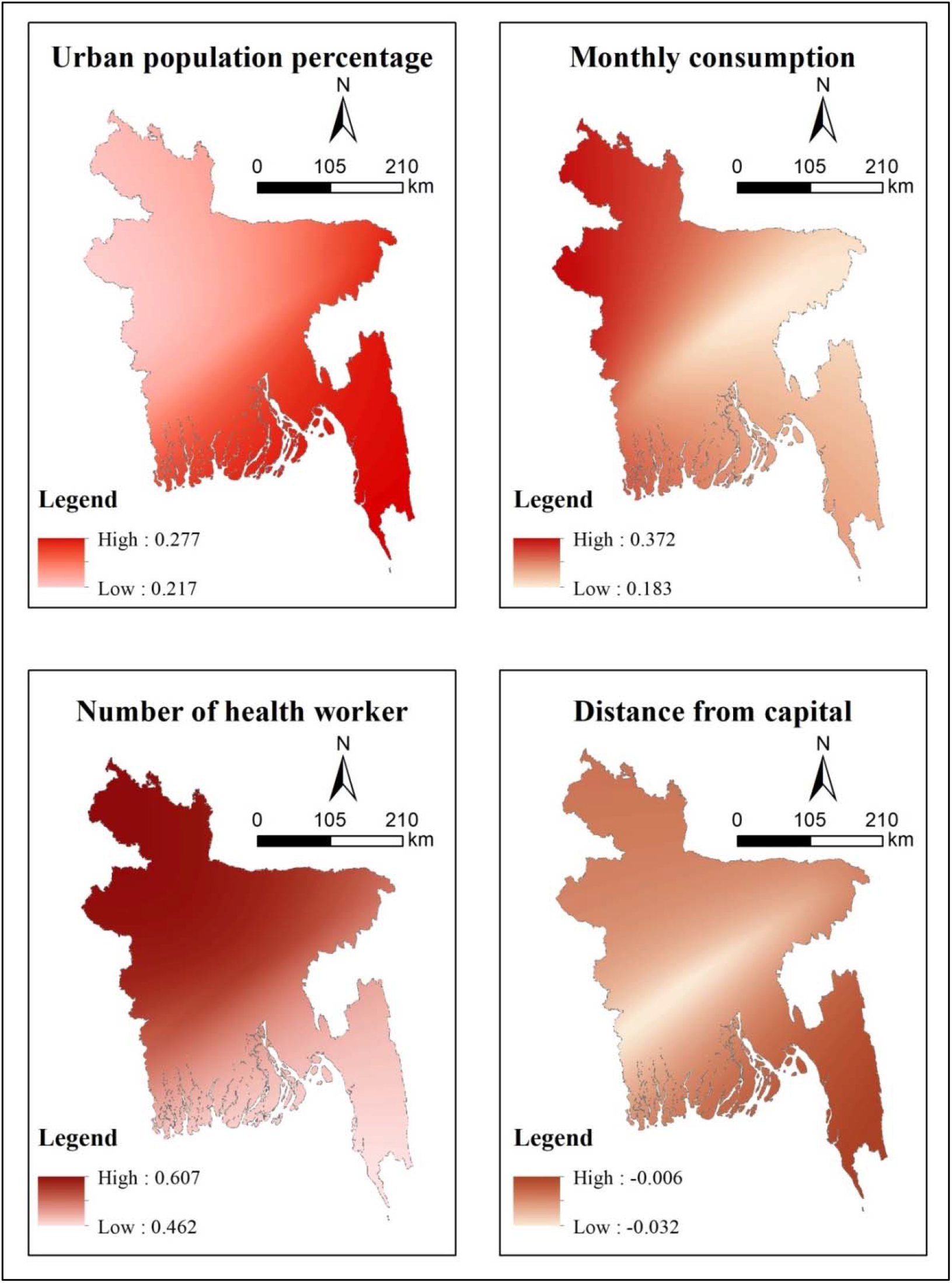
Spatial distribution of the coefficient values of urban population percentage, monthly consumption, number of health workers, and distance from the capital in describing COVID-19 incidence rates using GWR model

The results of mapping local *R^2^* values of the GWR model are demonstrated in **Figure 4**. Though decent local *R^2^* values were found for all districts, districts located in the southern parts of the country have comparatively lower local *R^2^* values than northern parts of the country, indicating a decent prediction of the model across the country especially in northern districts.

**Figure 4:**
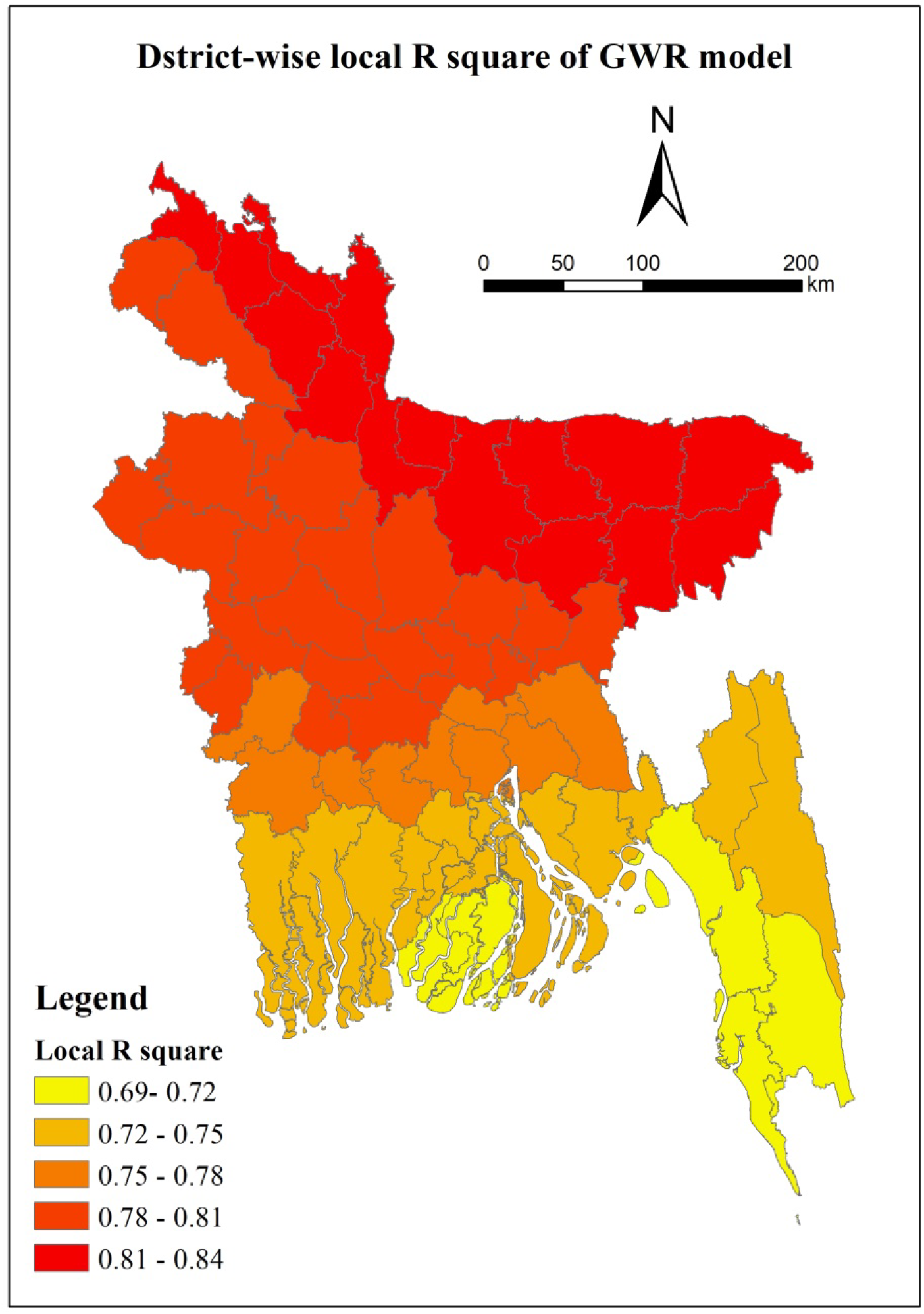
Spatial distribution of the local *R^2^* values of GWR model

## 4. Discussion

This study aimed to examine which factors have significant impact and substantial explanatory power to explain the COVID-19 incidence rates at district-level across Bangladesh with SRMs in GIS environment. These factors were grouped under six major themes: demographic, economic, weather, built environment, health, and community facilities. Results of the study showed that models with four variables– urban population percentage, monthly consumption, number of health workers, and distance from the capital– could explain relatively higher variation in the COVID-19 incidence rates in Bangladesh. Among the used four models, local SRM (GWR) was found to better explain the variation in COVID-19 incidence rates.

Urban population percentage was found to be proportionally related to COVID-19 incidence rates which implies urbanization paves the way to increase the likelihood of transmission of COVID-19. High urban population leads to increase movement and activities of people in high-density urban area (Hamidi et al., 2020; Ramírez-Aldana, Gomez-Verjan, & Bello-Chavolla, 2020). The higher the population density is, the more likelihood to come in proximate contact between an infector and infectee (You, Wu, & Guo, 2020). To be mentioned here, in this study, population density was also found strongly significant in univariate analysis and was not considered for developing overall model due to multicollinearity issue.

Households having higher monthly consumption have a tendency to purchase goods from commercially vibrant places which increases the potential of being affected by COVID-19 (You et al., 2020). Furthermore, higher consumption is associated with higher income level, lower poverty, as well as higher employment rates. Therefore, income and employment related activities trigger frequent travel and physical contact with people which could increase the risk of transmission of COVID-19 (Ehlert, 2020; Wheaton & Kinsella Thompson, 2020).

Number of health workers had a positive influence on the COVID-19 incidence rates as hospital acts as the hotspots of COVID-19 circulation. This result was found to be consistent with the findings of Mollalo et al. (2020). There was a severe shortage of personal protection equipment (PPE) in Bangladesh at the beginning of the pandemic. Therefore, a significant percentage (25%) of frontline health workers are obliged to tackle COVID-19 without any protection and thereby, a large number of health workers become infected by COVID-19 (Mahmud, 2020).

Distance from the capital was found inversely related to COVID-19 incidence rates. Dhaka, the capital and a megacity of Bangladesh, is the epicenter of pandemic in the country and has the most number of infected populations (**Figure 1**). In addition, Dhaka is the main commercial and administrative hub of the country; hence, it has a huge transport demand from different parts of the country. As, during this pandemic, public transport was under restriction across the country, districts in closer proximity to Dhaka, somehow, could manage to travel to this district more frequently than districts located further. These might be the reasons behind the result: the lower the distance of a district from the capital, the higher would be the likelihood of transmission of COVID-19.

Our study demonstrates that weather indicators had no significant influence on the occurrence rates of COVID-19. In literature, there are confounding findings regarding the influence of weather indicators on COVID-19 incidence: one group of researchers finds association between temperature or humidity and COVID-19 incidence (Ahmadi, Sharifi, Dorosti, Ghoushchi, & Ghanbari, 2020; Gupta, Raghuwanshi, & Chanda, 2020; Liu et al., 2020; Ma et al., 2020) while another finds no significance in this regard (Jüni et al., 2020; Mollalo et al., 2020; Xie & Zhu, 2020). There is a scope of rigorous and detailed study to understand the impact of weather related indicators on the occurrence of COVID-19.

This study showed that the impact of most of the community facilities, i.e., primary school, secondary school, college, growth center, rural market, religious establishment on COVID-19 incidence rates was insignificant as these facilities were shut down at the initial stage of COVID-19 pandemic. However, results of the univariate analysis shows that number of transit station and number of police station factors had profound influence to increase COVID-19 incidence. Though transit stations were also controlled through lockdown measures, stringent control was not implemented during festive and sudden opening and again closing decision of garments (Mamun, 2020). In addition, emergency activities, i.e., health care, food delivery, regular necessary goods marketing etc. were out of the scope of lockdown measures which also leaded to use transit stations. As a result, transit stations might become crowded which paved the way to transmit COVID-19. In addition, police had the responsibility to implement lockdown and social distancing in the field level during the pandemic. However, enough protection equipment was not available for the police officers to carry out their responsibility safely. In addition, overcrowding travel of police officers in a single police van made them more vulnerable to the virus (Javed, 2020).

One of the limitations of our study was data availability. Due to unavailability of individual or community level data, it is not logical to draw inferences at individual or community level. Another instance, weather data for 64 districts were interpolated from 28 station data in this study. The detailed weather related data availability might change the findings of the model. Another reservation is that spatial availability of COVID-19 testing center: in Bangladesh, there are few opportunities to test COVID-19 for people living in the remote area. Besides, there is a tendency among people not to test COVID-19 even though they have symptoms. Therefore, there might be an underestimation of COVID-19 cases. Furthermore, the influence of lockdown and other containment measures on COVID-19 incidence rates was not considered in this study. There is obvious to have variations in lockdown related policies and their implementation efficiency within a district. It might play an important role in the district-level to control the COVID-19 incidence rates but analyzing this influence would be out of the scope of this research.

## 5. Conclusion

Identification of possible determinants of virus transmission and spread is crucial, especially for coronavirus disease (COVID-19) which brought unprecedented shock globally. This study aimed to identify potential factors affecting the COVID-19 incidence rates at district-level across Bangladesh. To fulfill the aim, three global (OLS, SLM, and SEM) and one local (GWR) spatial models were developed in this study to identify potential demographic, economic, weather, built environment, health, and facilities related factors affecting the COVID-19 incidence rates.

First, univariate OLS models were developed where 17 factors were found statistically significant among the 32 considering factors. After that, a global multivariate OLS model was developed. The results of the model showed that four factors– urban population percentage, monthly consumption, number of health workers, and distance from the capital– were found to be statistically significant and had a *R^2^* value of 0.673. To address the issue of spatial dependency, SLM and SEM were further employed which resulted in increasing the *R^2^* values. Finally, a GWR model was developed to examine the spatial non-stationarity issue. The GWR model results showed improvement of model performance as explanatory powers increased to 78.6 % with the lowest AICc value (AICc = 340.49) compared to other models. Local *R^2^* values showed that model factors could better explain the COVID-19 incidence better in districts located northern part of the country compared to the southern regions. Theoretical investigations and empirical observations from this research offer an alternate view for the joint importance of the health and non-health determinants, which will help to develop policies aimed at preventing future epidemic crises.

## Data Availability

Data will be shared on request

